# PaiX Net: A Next-Generation Second-Opinion Platform for Pathology

**DOI:** 10.64898/2026.02.04.26345344

**Authors:** Jens Baumann, Bhavanikbhai Kanani, Shoeb Tamboli, Yuliya Kucherenko, Peter Fritz, Witali Aswolinskiy, Christoph Bosch, Martin Paulikat, John K.L. Wong, Bharti Arora, Myroslav Zapukhlyak, Jens Eickmeyer, Marina Pavlova, Roman Laskorunskyi, Yulia Kindruk, Simon Kalteis, Nishan Tamang, Manasi Aichmüller-Ratnaparkhe, Gizem Yazli, Gizem Uluc, Patrick Adam, Danny Quick, Christian Aichmüller

## Abstract

Pathology faces persistent challenges including a global shortage of specialists, uneven access to expertise, increasing diagnostic complexity, and a growing need for second-opinion consultations. While digital and telepathology platforms address parts of this problem, existing solutions often trade accessibility for structured, workflow-aware clinical integration. At the same time, multimodal medical AI shows promise for diagnostic support but raises concerns regarding transparency, automation bias, and clinical accountability.

We present **PaiX Net**, a structured, AI-augmented second-opinion platform designed to support collaborative pathology consultation while preserving human decision ownership. The platform integrates standardized case templates, moderated expert discussion, and human-centered AI assistance within a scalable, browser-based architecture compliant with data protection requirements. AI support is embedded at defined workflow stages to assist with case structuring, summarization, and exploratory interpretation, while diagnostic conclusions remain under expert control. To mitigate automation bias, AI-generated content is visually separated, collapsed by default, and presented only after independent expert input.

PaiX Net incorporates a multimodal medical AI model (MedGemma-4B), selected for its open availability and computational efficiency, and fine-tuned on curated, anonymized consultation cases. An illustrative retrospective evaluation demonstrates substantial reductions in case preparation time and modest but consistent improvements in diagnostically relevant summaries.

PaiX Net illustrates how structured, human-centered AI integration can enhance access to expert second opinions while maintaining clinical accountability and supporting continuous human–AI learning in digital pathology.

## 1. Introduction

Pathology plays a central role in modern medicine, underpinning accurate diagnosis, prognosis, and treatment planning [1]. However, there is a growing global shortage of pathologists, which has a direct impact on healthcare delivery and diagnostic response times [2]. This shortage is further compounded by an uneven distribution of expertise between global regions, as well as the increasing complexity of pathological assessments driven by advances in molecular diagnostics and digital imaging. Especially the time required for specialized education, for example, 11 years or more in the USA [3], makes clear that addressing this shortage will likely take decades rather than years. These challenges result in diagnostic delays, variability in interpretation, and limited access to expert opinions, particularly in low-resource or rural settings [4, 5]. In addition, studies have demonstrated that even in settings with good availability of pathologists, obtaining a second-opinion can substantially improve diagnostic accuracy and patient outcomes [6, 7, 8, 9].

However, the opportunities for such consultations remain constrained by logistical, institutional, and geographic barriers.

We identify four interrelated systemic challenges:

1. an overall shortage of trained experts,
2. an unequal geographic and institutional distribution of expertise,
3. the high level of experience required for reliable diagnostic decision-making,
4. and the frequent need for second-opinions, particularly in complex or ambiguous cases.

While the long-term workforce shortage (1) can only be addressed through sustained global investment in training, the short- and medium-term challenges (2–4) require more efficient use of existing expertise. Telepathology/Digital pathology platforms can offer a unifying approach to all four challenges by enabling remote collaboration, feedback, knowledge transfer, and collective learning across institutional and geographic boundaries.

### Scope and contributions

This manuscript is primarily a *design, architecture, and implementation* paper describing a second-opinion platform for digital pathology and its human-centered integration of AI-assisted support. We complement this with an ecosystem-mapping (scoping) analysis of existing second-opinion mechanisms and an *illustrative* retrospective evaluation of the embedded model to contextualize feasibility. Accordingly, the presented results should be interpreted as evidence of technical plausibility and workflow fit rather than as a full clinical validation study.

## 2. Background and Motivation

Advances in communication technology and medical imaging have enabled telemedicine and, in particular, telepathology as a means of remotely observing and interacting with tissue specimens. Telepathology is commonly categorized into static and dynamic approaches. Static telepathology involves the capture and asynchronous transmission of selected regions of interest (ROIs) for later review, whereas dynamic telepathology enables real-time interaction with specimens, either via robotic microscope control or synchronous video-based modalities [10, 11]. Whole-slide imaging (WSI) occupies an intermediate position, providing a complete high-resolution digital representation of the tissue while still supporting interactive navigation and inspection.

### Identifying Platforms

To identify which platforms are currently relevant for obtaining second-opinions in digital pathology, we conducted both an academic literature search and a systematic web search. This process can be characterized as a scoping review aimed at mapping the landscape of platforms used for pathology second-opinions, rather than a systematic review of outcomes or performance. Accordingly, inclusion criteria emphasized functional relevance and real-world usage over formal validation.

For the literature search (Figure 1), we queried PubMed using the following terms: `((digital pathology) DR (telepathology)) AND ((forum) DR (platform)) AND (second opinion)`.

**Figure 1.**
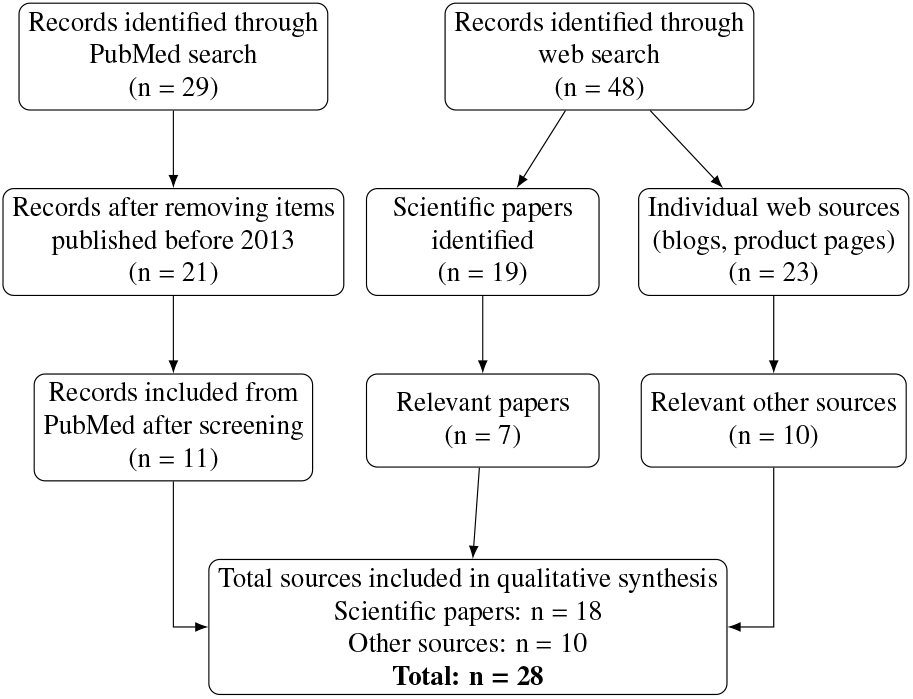
Flow diagram of the platform identification process.

For the web search, we screened the first page of Google results using the queries: `digital pathology second opinion platform, digital pathology second opinion forum, telepathology pathology second opinion platform` and `telepathology second opinion forum.`

We defined “second-opinion pathology platforms” as online tools that support case-based discussion of pathology among pathologists and/or trainees, irrespective of whether their primary focus is education, diagnostics, or consultation.

From PubMed, we initially identified 29 papers. We excluded papers published before 2013, resulting in 21 remaining articles, of which 11 described or evaluated a platform for sharing second-opinions.

The web search yielded 48 relevant results, including 19 scientific papers. Seven of these papers were not identified through the PubMed query and were therefore added to the 11 publications above, resulting in a total of 18 papers.

In addition, the web search identified 10 non-scientific sources, such as blog posts or product introductions, which provided further insight into currently available platforms.

Across these sources, the identified platforms can be grouped into public and private communication channels, general-purpose data-sharing tools, and dedicated clinical or institutional data-sharing platforms (see Table 1).

**Table 1:** Grouping of identified platforms used for pathology case exchange and second-opinion consultation, categorized by communication modality and level of clinical integration.

| Group | Platforms |
| --- | --- |
| Public channels | YouTube, Twitch [12]; Facebook, Twitter/X [13] |
| Private channels | Phone [14]; Email (commonly used informally for sharing images, questions or platform notifications); WhatsApp, Telegram, LINE [13]; Zoom [15, 16]; Skype [11] |
| Datasharing tools | Box [15]; case.io [17]; Google Drive [13] |
| Clinical/ Institutional | iPath [18, 19, 20, 21, 22, 11]; mPathology [23, 24]; dPathology [25]; HyPathology [26]; PathPresenter [16]; UPMC [27, 28]; PIE [29]; scanner software [14, 30, 31]; navify [32]; tribun [33]; PathoZoom [34]; custom setups [35, 36] |

The platforms identified above collectively form a heterogeneous ecosystem for pathology case exchange (Table 1), spanning public social media, private communication tools, generic cloud-based data-sharing services, and dedicated clinical or institutional telepathology systems. Each category supports distinct workflows and levels of structure, which strongly influence how cases are shared, discussed, and archived.

### Public communication channels

Public platforms such as X (the platform formerly known as Twitter), YouTube, Twitch, Facebook, Instagram, and Reddit enable rapid, low-barrier distribution of cases and opinions. Communities including #PathTwitter/#PathX illustrate how pathologists and trainees use short-form posts, hashtags, and representative images to solicit feedback, debate diagnoses, and share teaching material at scale [13, 37, 38, 39, 40]. Video-centric platforms further support explanatory content, annotated slide reviews, and live case discussions [12]. However, these platforms encode cases almost exclusively as free text accompanied by one or a few compressed images, corresponding to an extreme form of *static telepathology*, with little or no standardized metadata regarding specimen type, stain, anatomical site, or diagnostic context. As a result, systematic retrieval, longitudinal follow-up, and integration with laboratory information systems are not feasible. While effective for rapid peer engagement and informal learning, these platforms implicitly shift diagnostic reasoning from a case-centered to a post-centered paradigm, constraining depth, reproducibility, and clinical accountability.

### Private communication channels

Private tools, including phone calls, email, instant messaging applications (e.g., WhatsApp, Telegram, LINE), and video-conferencing platforms such as Zoom or Skype, remain central to day-to-day consultation and urgent decision-making [14, 13, 15, 16, 11]. Their strengths lie in immediacy, trust, and conversational flexibility, making them particularly suitable for frozen sections or time-critical second-opinions, and aligning closely with *dynamic telepathology* through real-time visual and verbal interaction. Nevertheless, these exchanges are typically ephemeral, poorly structured, and confined to personal networks. Shared images are often screenshots or smartphone photographs detached from standardized identifiers, effectively reverting dynamic interactions back to unstructured *static* artifacts and limiting reproducibility, auditability, and secondary reuse.

### General-purpose data-sharing tools

Cloud-based storage and case-sharing services such as Box, Google Drive, and case.io provide higher-resolution image sharing and asynchronous access [15, 17, 13]. While these tools offer basic access control and persistence, and can support larger whole-slide image–based *hybrid telepathology* workflows, they are not pathology-native and lack structured schemas for diagnostic metadata, uncertainty, or outcome annotation. Consequently, clinical meaning remains implicit and external to the platform itself. As a result, essential diagnostic semantics, such as uncertainty, differential diagnoses, and clinical follow-up, remain external to the platform and inaccessible to computational or longitudinal analysis.

### Clinical and institutional platforms

Dedicated telepathology systems, including iPath, mPathology, dPathology, HyPathology, PathPresenter, PIE, UPMC-hosted solutions, scanner-integrated software, and commercial enterprise platforms such as navify, tribun, and PathoZoom, are explicitly designed for clinical workflows [18, 19, 20, 21, 22, 23, 24, 26, 16, 27, 29]. These systems typically support whole-slide image uploads, structured case descriptors, traceable consultations, and regulated access, thereby enabling *hybrid telepathology* workflows that combine static whole-slide review with dynamic consultation, and are often embedded within institutional or national networks. While technically robust, they tend to prioritize compliance, governance, and local workflows, which may limit openness, cross-institutional reuse, or exploratory data-driven analyses.

### Comparative perspective

Taken together, these platform categories reveal a fundamental trade-off between accessibility and structure. Public and private communication channels maximize reach, responsiveness, and informal peer engagement, relying predominantly on static and dynamic telepathology, respectively, and enabling rapid access to collective expertise across geographic boundaries, but at the cost of standardization, traceability, and integration with clinical workflows. Conversely, institutional telepathology systems emphasize hybrid telepathology by integrating static whole-slide imaging with dynamic consultation, providing structured, auditable case representations and regulatory compliance, but often imposing substantially higher barriers to participation. These barriers include institutional enrollment, contractual agreements, technical integration with local infrastructure, and limited support for spontaneous or cross-institutional consultations, which can restrict participation by smaller centers, trainees, and pathologists in low-resource settings. As a result, access to expert second-opinions remains unevenly distributed despite the availability of technically mature systems. General-purpose data-sharing tools occupy an intermediate position, offering persistence and basic access control, but lacking pathology-specific semantics, governance, and workflow support.

This fragmentation has practical consequences beyond daily consultation. Recent large-scale vision–language datasets for pathology increasingly draw on heterogeneous and opportunistic data sources, including social media posts and loosely curated repositories [41, 42, 43]. Although substantial in scale, such datasets frequently lack consistent diagnostic labels, standardized metadata, and complete clinical context, which limits their suitability for supporting expert-level diagnostic reasoning or downstream clinical integration. Collectively, these observations highlight the need for platforms that preserve the structured, workflow-aware foundations of established telepathology systems while lowering barriers to participation and improving data organization, interoperability, and analytical reuse.

## 3. PaiX Net: System Design and Workflow

PaiX Net is a structured, AI-augmented platform designed to support collaborative second-opinion consultations in pathology. The platform integrates standardized case representation, moderated expert discussion, and human-centered AI assistance within a unified, workflow-aware environment. Building on established asynchronous case-based telepathology platforms, most notably iPath, PaiX Net extends these paradigms to support contemporary digital pathology workflows and continuous human–AI knowledge exchange (Figure 2).

**Figure 2.**
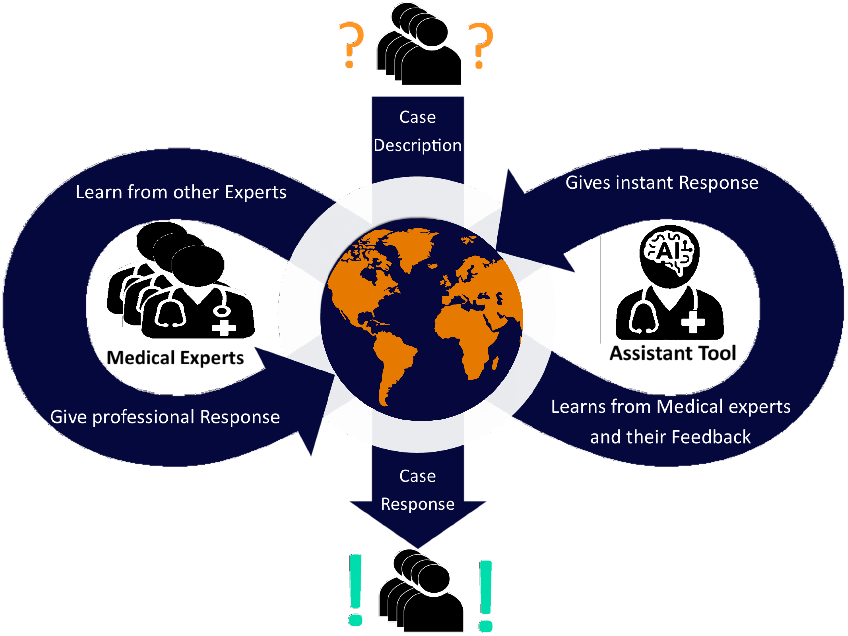
Conceptual overview of PaiX Net. The platform combines expert-driven case discussion with AI-assisted analysis to support high-quality second-opinion pathology. Human experts and AI models interact in a continuous feed-back loop, enabling diagnostic support, peer learning, and supervised model improvement based on accumulated case knowledge.

### 3.1. Design Objectives and Method Overview

Second-opinion pathology involves heterogeneous case material, limited availability of subspecialty expertise, and strong requirements for diagnostic accountability. In practice, existing solutions emphasize different design priorities, ranging from rapid, low-barrier exchange to structured, traceable workflows. PaiX Net is designed to address this gap by explicitly combining open participation with structured, workflow-aware case representation.

The platform is guided by five design objectives: (i) structured and reusable case representation; (ii) low-barrier, cross-institutional participation; (iii) moderated and traceable expert discussion; (iv) human-centered AI assistance; and (v) explicit mitigation of cognitive and automation bias.

Operationally, PaiX Net structures second-opinion consultation as a multi-stage workflow comprising case submission, expert interpretation, moderated discussion, consensus formation, and knowledge capture. AI assistance is integrated as a supportive component at defined stages, while all diagnostic decisions remain under human oversight.

### 3.2. System Architecture

PaiX Net follows a modular, service-oriented architecture that separates user interaction, data management, and AI services. This separation supports scalability, regulatory compliance, and the independent evolution of AI components without disrupting established clinical workflows or governance structures.

As illustrated in Figure 3, the architecture consists of three logical tiers: (i) an application tier managing user interaction, workflow orchestration, and moderation; (ii) a data tier for structured case data and image storage; and (iii) an AI services tier providing automated case structuring and interpretive assistance. All interactions occur via encrypted communication channels, ensuring data protection and operational reliability.

**Figure 3.**
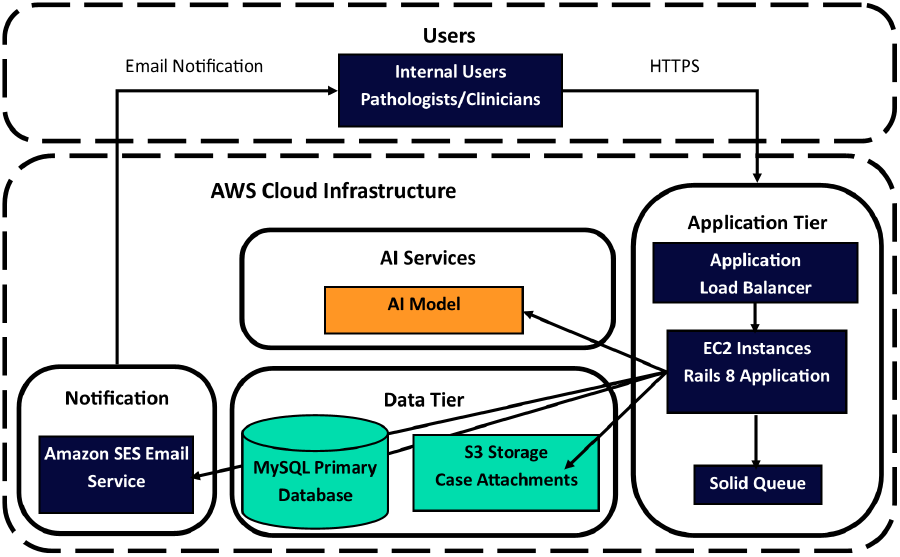
System architecture of the PaiX Net platform, illustrating its secure and scalable deployment within the AWS cloud environment. The platform is organized into three core tiers: (i) the Application Tier, where a Rails-based web application runs on EC2 instances behind an Application Load Balancer and manages asynchronous tasks via Solid Queue; (ii) the Data Tier, comprising a MySQL primary database and S3 object storage for case attachments; and (iii) the AI Services, which hosts the AI model responsible for automated case structuring and assistance. Integrated notification services such as Amazon SES enable automated email alerts to clinicians and pathologists. All user interactions occur via HTTPS, ensuring end-to-end data protection and operational reliability.

### 3.3. Case Representation and Consultation Workflow

Cases are submitted using structured templates capturing patient history, diagnostic questions, imaging data, and supporting materials. Subspecialty-specific templates dynamically expose relevant fields, improving completeness and standardization (Figure 4).

**Figure 4.**
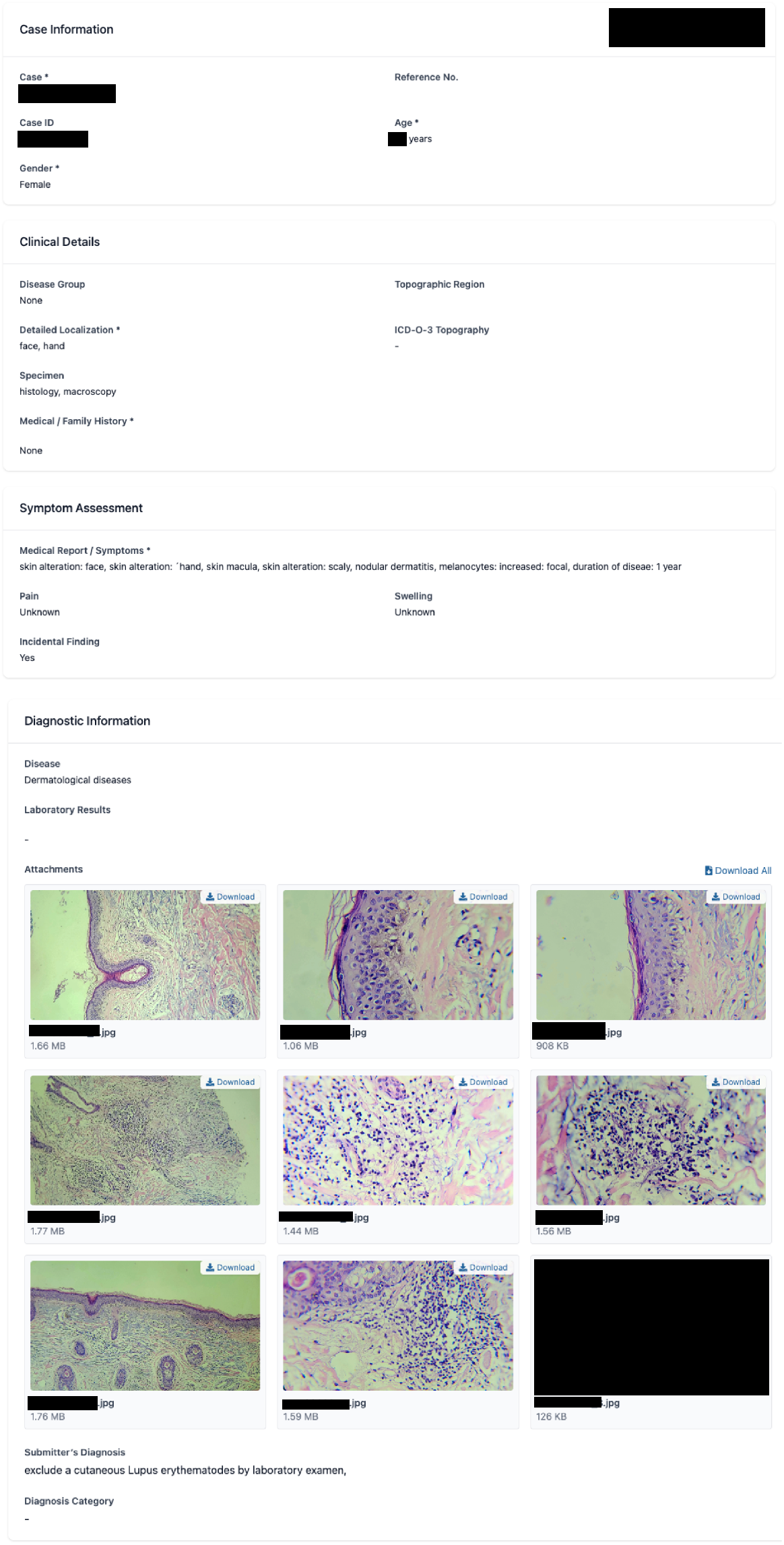
Completed case information template showing clinical details, symptom assessment, diagnostic information, and attached images including histology slides and a clinical photograph.

Cases progress through a lifecycle with defined states (*open, solved, unsolved*). Expert consensus enables formal case closure, and supporting tools allow standardized coding and export of case materials. This structure ensures traceability of diagnostic reasoning, supports longitudinal reuse of consultation data, and enables secondary analysis under appropriate governance.

### 3.4. Expert Interaction and Moderation

Consultations are organized within topic-specific groups moderated by designated experts. Moderators manage access permissions, approve membership requests, and ensure thematic coherence. Role-based permissions enable structured oversight.

Each case includes a persistent, threaded discussion environment supporting focused, evidence-based dialogue. Moderators can highlight authoritative contributions and formally close cases with an explicit diagnostic outcome. To mitigate anchoring and authority bias, the platform supports mechanisms such as delayed visibility of existing comments until users submit their own initial assessment.

### 3.5. AI-Augmented Support and Human–AI Feedback

AI assistance is integrated at two points in the workflow. During case creation, AI supports extraction of structured information from unstructured inputs such as photographs of reports and prompts users for missing data. Additionally, the *ICD-O-3 Topography* code is generated in this step. After submission, the AI generates an exploratory summary of the provided information, highlighting key findings, additional potentially relevant observations, and suggested next steps (Figure 5).

**Figure 5.**
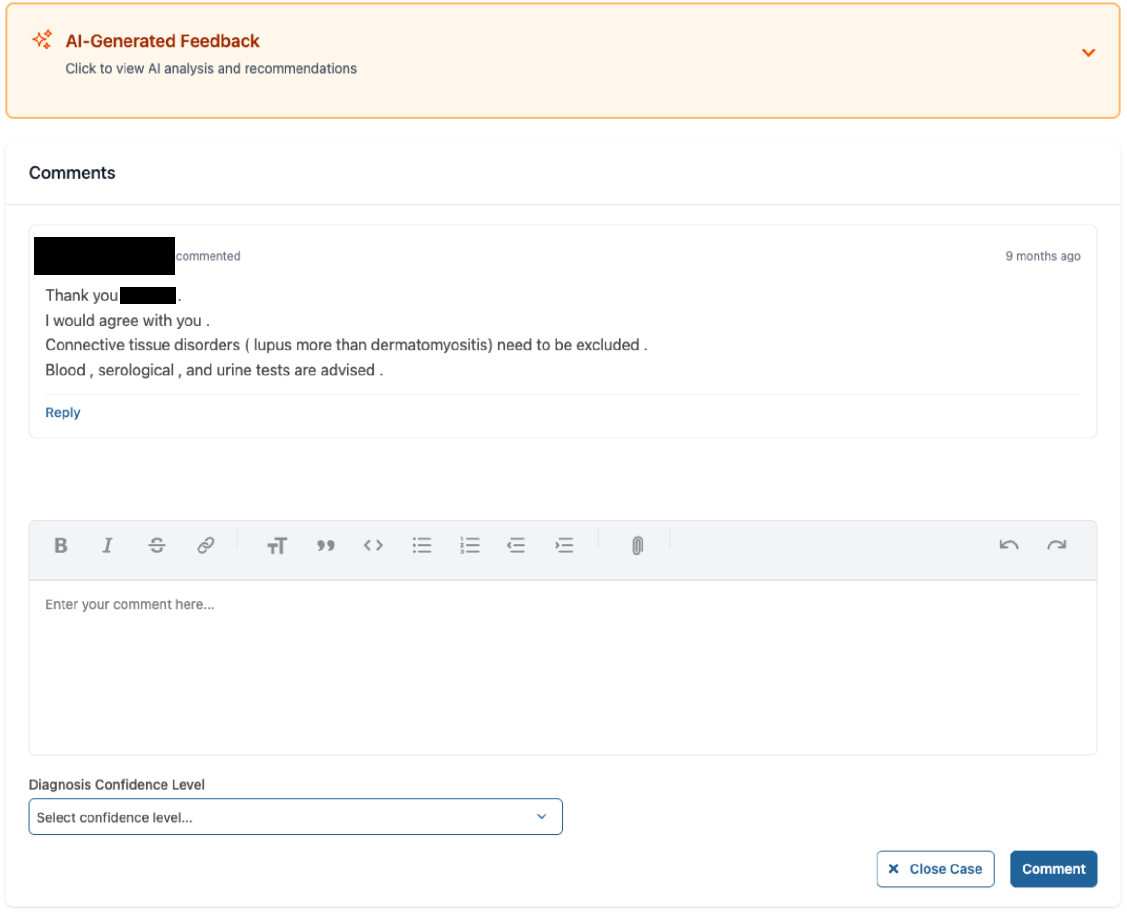
Case discussion section showing AI response and expert comments. Comments can address the main case or reply to previous comments. The AI section (red) is collapsed by default to distinguish it from human responses.

To mitigate automation bias, AI-generated content is visually separated from human discussion and collapsed by default. Users are encouraged to provide independent assessments prior to reviewing AI output. Expert feedback on AI-generated content is systematically captured and used to inform iterative model refinement, establishing a continuous human–AI feed-back loop while preserving human oversight (Figure 6).

**Figure 6.**
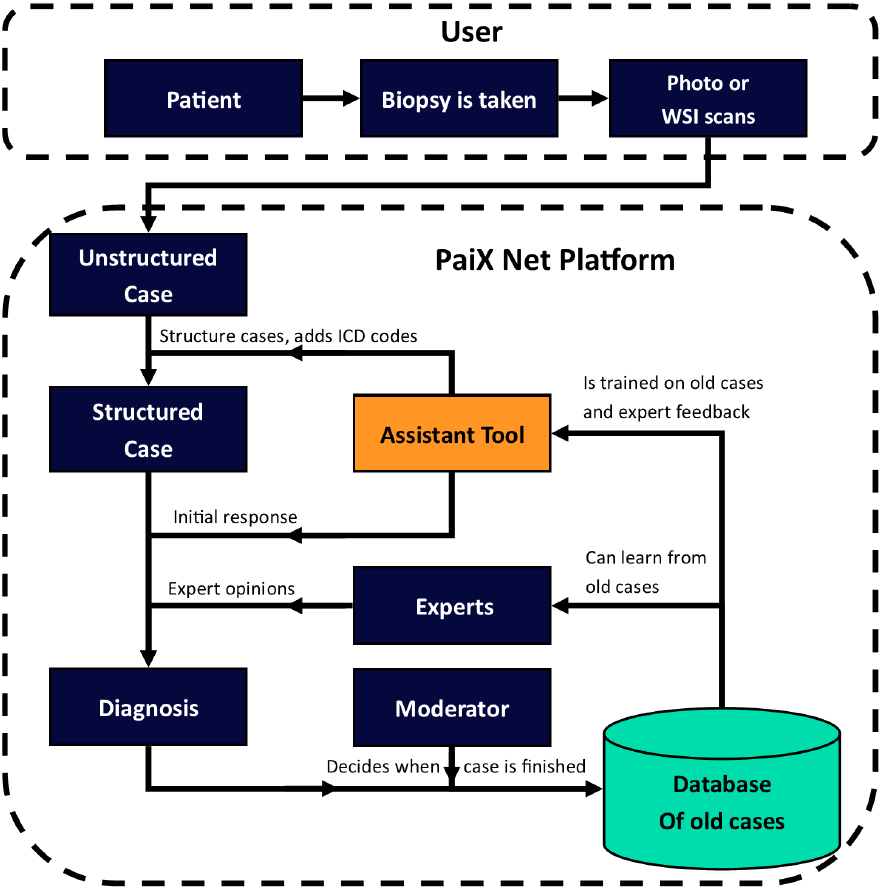
Alternative conceptual architecture illustrating human–AI knowledge flow in PaiX Net. The diagram highlights the interaction between unstructured case inputs (text and images), AI-assisted case structuring, expert interpretation, moderated discussion, and final diagnostic decisions. Expert feedback is continuously captured and stored, enabling supervised learning and iterative improvement of the AI assistant while preserving human oversight and clinical accountability.

For the AI model, MedGemma-4B, an instruction-tuned multimodal model released under Google’s Health AI Developer Foundations program [44] was selected due to its balance of medical domain competence, multimodal capability, computational efficiency, and open availability. The model combines a SigLIP-based image encoder [45] with a 4-billion-parameter language backbone pretrained on large-scale, medical datasets. Compared to generic vision–language models, MedGemma-4B demonstrates substantially improved performance on medical benchmarks, including histopathology-related tasks, while maintaining a manageable computational footprint suitable for low-latency inference and fine-tuning on consumer-grade hardware. These characteristics align well with the practical constraints of interactive, expert-in-the-loop second-opinion workflows. To further adapt the model to the PaiX Net use case, MedGemma-4B was fine-tuned on a curated dataset of anonymized pathology consultation cases. This fine-tuning focused on improving multi-image interpretation, summarization of salient findings, and generation of structured outputs compatible with the platform’s case templates.

### 3.6. Implementation and Governance

PaiX Net is implemented as a browser-accessible web application that does not require specialized client software. Data handling processes are designed to comply with applicable data protection regulations, including GDPR. Role-based access control, and moderated governance ensure appropriate clinical accountability and traceability.

## 4. Evaluation

The evaluation of PaiX Net is designed as an illustrative feasibility assessment rather than a definitive clinical validation. The objective is to assess workflow efficiency, platform responsiveness, and the practical utility of AI-assisted case interpretation within real-world second-opinion pathology scenarios. Accordingly, results should be interpreted as evidence of technical plausibility and workflow fit, not as a measure of standalone diagnostic performance.

### 4.1. Efficiency of AI-Assisted Case Creation

To assess workflow efficiency, the time required for structured case creation was measured with and without AI assistance. For users familiar with both the platform and the respective cases, manual case entry required an average of 2 minutes and 41 seconds. When using the AI-assisted case creator, the total time was reduced to an average of 53 seconds. This duration includes approximately 20 seconds of automated processing time and 33 seconds for user review and completion.

### 4.2. Consultation Turnaround Times

Platform data from the iPath forum for the year 2023 was analyzed to characterize expert response times. The year 2023 was selected because it is recent enough to ensure continued relevance, while also being sufficiently distant to guarantee that all included cases have reached completion.

Across 6,119 cases, the median time from case submission to the first expert response was approximately 2.5 hours. Within 21 hours, 90% of cases had received at least one reply. The median time to a final expert response was 10 hours, while approximately 90% of cases reached a final response within three days.

For cases requiring additional input from the original poster, the median time to a final expert comment increased to over one day.

### 4.3. AI-Assisted Interpretation Performance

To assess the diagnostic relevance of the AI assistant, model-generated interpretations were compared against original diagnoses and expert review. Table 2 summarizes the results for the pretrained MedGemma-4B model and the fine-tuned variant deployed within PaiX Net. Fine-tuning resulted in an improvement, increasing the number of correct interpretations from 137 to 146 cases.

**Table 2:** Performance comparison of the pretrained MedGemma-4B model and its fine-tuned variant on 285 historical consultation cases. Model predictions were evaluated by expert pathologists and categorized as correct, partially correct, or not correct. “Partially correct” indicates diagnoses that captured relevant tissue features or were overly broad but led to an incorrect or incomplete conclusion.

| Model | Not correct | Partially correct | Correct |
| --- | --- | --- | --- |
| MedGemma-4B | 112 (39.30%) | 36 (12.63%) | 137 (48.07%) |
| Fine-tuned | 107 (37.54%) | 32 (11.23%) | **146 (51.23%)** |

Qualitative expert review identified several recurring failure modes. These included diagnostically rare entities, borderline cases with limited contextual information, and scenarios requiring nuanced integration of clinical history.

## 5. Discussion

The findings from Subsection 4.1 demonstrate that AI-assisted case structuring can substantially reduce the time required for standardized case submission while preserving user oversight and content accuracy. Although the AI-assisted workflow introduces an average idle time of approximately 20 seconds during automated processing, this delay can be mitigated by overlapping the processing phase with image upload, further improving overall efficiency.

Results from Subsection 4.2 indicate that initial expert responses are typically provided rapidly, often within a few hours. However, reaching a definitive conclusion frequently requires substantially more time and the involvement of multiple participants. In particular, cases in which the initial response highlights missing clinical information or recommends additional diagnostic tests tend to experience extended turnaround times.

This delay is largely attributable to the asynchronous nature of forum-based consultations. Original posters must wait for expert feedback, experts then wait for additional information, and subsequent expert responses are again delayed. This iterative exchange introduces significant latency into the diagnostic process. These observations underscore the potential value of an instant, AI-assisted response at the time of case submission, helping users identify missing information early and reduce avoidable delays.

The fine-tuned model (Subsection 4.3) demonstrates a modest but consistent improvement over the pretrained baseline, particularly in generating diagnostically relevant summaries and structured interpretations. Despite this improvement, performance remains heterogeneous across subspecialties and case complexities, reflecting the inherent challenges of multimodal medical reasoning. Importantly, these gains indicate that targeted fine-tuning on structured, expert-annotated consultation data can enhance the practical utility of multimodal AI models in second-opinion support settings. Accordingly, these findings motivate a prospective, iterative development strategy in which continuous expert feedback collected within PaiX Net is leveraged to further refine model behavior and adapt to real-world diagnostic workflows.

Rather than positioning the AI component as a diagnostic authority, PaiX Net explicitly frames AI output as exploratory and supportive. Design choices such as visual separation, collapsed default presentation, and explicit encouragement of independent expert assessment are intentionally employed to mitigate automation bias. Within this configuration, AI assistance complements human expertise while preserving clinical accountability and decision ownership.

## 6. Summary and Conclusions

Pathology faces a confluence of systemic challenges, including a global shortage of trained experts, unequal access to specialized expertise, increasing diagnostic complexity, and a persistent need for second-opinions in difficult or ambiguous cases. While long-term solutions require sustained investment in education and workforce development, short- and medium-term improvements depend on more effective use of existing expertise and better mechanisms for collaboration across institutional and geographic boundaries.

In this work, we surveyed the current ecosystem of platforms used for pathology second-opinion exchange, ranging from public social media channels and private communication tools to general-purpose data-sharing services and dedicated clinical telepathology systems. Our analysis highlights a persistent trade-off between accessibility and structural rigor in existing second-opinion platforms, which informed the design decisions underlying PaiX Net.

To address these gaps, we introduced **PaiX Net**, an AI-augmented second-opinion platform designed to balance openness with structure. Building on the conceptual foundations of earlier telepathology systems such as iPath, PaiX Net integrates structured case workflows, moderated expert discussion, and AI-assisted support within a scalable, privacy-preserving web-based architecture. The platform explicitly supports collaborative intelligence by enabling bidirectional knowledge transfer between human experts and AI models. Human oversight ensures transparency, mitigates automation bias, and preserves clinical accountability, while AI assistance supports case structuring, summarization, and exploration of differential diagnoses.

A key technical component of PaiX Net is the integration of a multimodal medical AI model, MedGemma-4B, selected for its balance of efficiency, domain-specific competence, open availability, and structured output capabilities. Fine-tuning on curated consultation cases further aligns the model with real-world pathology workflows. Importantly, AI-generated content is intentionally separated from human discussion and designed to support, rather than steer, expert reasoning.

Overall, PaiX Net demonstrates how structured, AI-augmented platforms can lower barriers to expert consultation while preserving the rigor, traceability, and interoperability required for clinical and analytical reuse. By combining global expert participation with workflow-aware digital infrastructure, PaiX Net has the potential to improve diagnostic consistency, accelerate access to second-opinions, and create a growing, structured corpus of expert-annotated cases. Such platforms represent a critical step toward more equitable access to pathology expertise and provide a foundation for future human-centered, data-driven advances in diagnostic medicine.

## Data Availability

The data used in this study are not publicly available due to institutional, ethical, and privacy constraints.

## Code and Data Availability

The code and data used in this study are not publicly available due to institutional, ethical, and privacy constraints.

## Ethics Approval and Consent to Participate

This study was conducted as a retrospective analysis of anonymized pathology consultation data obtained from iPath. All data was anonymized at source prior to transfer, and no identifiable patient information was accessed by the research team. The study was reviewed by Paicon’s Data Protection Officer, which determined that formal ethics approval and informed consent were not required due to the retrospective design and exclusive use of anonymized data. All data handling was conducted in accordance with GDPR.

